# Evaluation of Turkish social distancing measures on the spread of COVID-19

**DOI:** 10.1101/2020.04.28.20083550

**Authors:** İrfan Civcir

## Abstract

The coronavirus disease (COVID-19) affecting across the globe. The government of different countries has adopted various policies to contain this epidemic and the most common were social distancing and lockdown. We use a simple log-linear model with intercept and trend break to evaluate whether the measures are effective preventing/slowing down the spread of the disease in Turkey. We estimate the model parameters from the Johns Hopkins University (2020) epidemic data between 15th March and 16th April 2020. Our analysis revealed that the measures can slow down the outbreak. We can reduce the epidemic size and prolong the time to arrive at the epidemic peak by seriously following the measures suggested by the authorities.

## 1. Introduction

With the spread of the COVID-19, governments faced with the serious problem of how best to save people’s lives and minimize long-term economic damage. Because of the absence of pharmaceutical intervention, the most effective available interventions for national and global control and mitigation of COVID-19 is social distancing to reduce the transmission of COVID-19. The government of different countries is taking measures to contain this epidemic and the most common are travel restrictions, isolation of confirmed cases, and quarantine of exposed persons, curfews, school closures, banning of mass gatherings, social distancing, mandatory wearing of masks, isolation of ill persons, and appropriate disinfection and/or hygiene measures (Cowling and Aiello, 2020).

Like all other countries, Turkey also started taking measures to fight against the rapid spread of COVID-19. Several measures are taken in response to the evolving novel Coronavirus disease (COVID-19). These are movement restrictions, social distancing, and public health measures. On 11 March Turkey’s first confirmed coronavirus case was announced and on 12 March 2020, the Turkish government announced critical social distancing measures that primary schools, middle schools, high schools and universities, public libraries, bars, night clubs, and discotheques would be closed starting from 16 March 2020. On 16 March 2020, meetings in all closed and open prisons are suspended, prayer gatherings in mosques are banned nationwide, all public gathering places such as cafes, movie theatres, gyms are closed except shops and restaurants not offering music. On 19 March, sports matches also postponed. On 20 March, all kinds of cultural and scientific activities/meetings were suspended. Free public transportation for elderly people was temporarily suspended in several metropolitan cities.

On 21 March, the activities of barbershops, hairdressers, and beauty salons were stopped. Flights to in a total of 68 countries had stopped. A total curfew announced for those who are over the age of 65 or chronically ill. Restaurants, dining places, and patisseries were to be closed to the public for dining in and were only allowed to offer home delivery and take-away. On 27 March, all international flights were terminated, and intercity travel was subject to approval by the state governors. Furthermore, historical sites and picnic areas are closed on weekends. On 3 April 2020, the government issued a 15-day entry ban to 30 metropolitan cities as well as Zonguldak, which is extended later. Furthermore, the curfew was extended to people younger than 20 years old. Using masks in public places became mandatory (https://en.wikipedia.org/wiki/2020_coronavirus_pandemic_in_Turkey).

This article aims to determine the effectiveness of government measures to prevent or slow down the spread of Covid-19 in Turkey. We would anticipate the consequences of these measures to be visible as of 21 March 2020 and beyond. As expected, if these policy measures work to prevent or slow down the spread of Covid-19, the growth rate of the number of confirmed cases should display a slowdown starting from around 21 March.

The structure of the paper is as follows. Section 2 describes the data for confirmed cases of Covid-19 in Turkey. In Section 3, a simple model set up to determine the average growth rate of log cumulative confirmed cases, and this model is tested for intercept and trend breaks. In Section 4, the estimation results are presented and discussed. Section 4 concludes.

## 2. The data

To understand the effects of the government measures on the spread of Covid-19 in Turkey, we investigate the development of the number of confirmed cases of Covid-19 in Turkey between 15 March and 26 April 2020.

There are several sources of data. Official data can be obtained from the Ministry of Health (2020). Other main alternative sources are the World Health Organisation, the European Centre for Disease Prevention and Control and the Johns Hopkins University (2020). We used the Johns Hopkins University (2020) data, because it links data from the Health Ministry, the World Health Organization.

There are slight outliers at the beginning of the sample, where the case number is low but the variation is relatively high, therefore, we started our investigation after 10 or more cases reported which start from 15 March. Between 15–20 March, the growth rate of cumulative confirmed cases is about 123%, which implies that a doubling of confirmed cases every 0.57 days. Between 21–26 March, the growth rate of the confirmed cases drops to 33%, implying that the doubling of confirmed cases every 2.13 days. Between 27 March and 1 April, the average growth rate dropped to 28%, and the doubling of confirmed cases improved to every 2.47 days. Between 2–11 April, the growth rate dropped to 13 percent, 12–19 April to 8 percent, and 20–26 April to 5 percent and the doubling of the confirmed cases improved to 14.5 days, which is in line with the lagged impact of the policy measures taken by the Turkish authorities.

Figure 1 plots the Covid-19 cumulative confirmed cases in levels and logs from 15 March to 26 April. As can be seen from the levels figure on the left side, the cumulative confirmed Covid-19 cases increasing exponentially, taking logs of this variable yields a time series with a linear trend. These figures indicate that a linear time trend may characterize the dynamics of log confirmed cases well, although there are some levels and trend shifts in the sample. There are a few outliers at the beginning of the sample. In the first week (12–18 March), an outbreak of Covid-19 among returning travelers from Europe, the US, Iran, and Saudi Arabia increased the case numbers.

**Figure 1:**
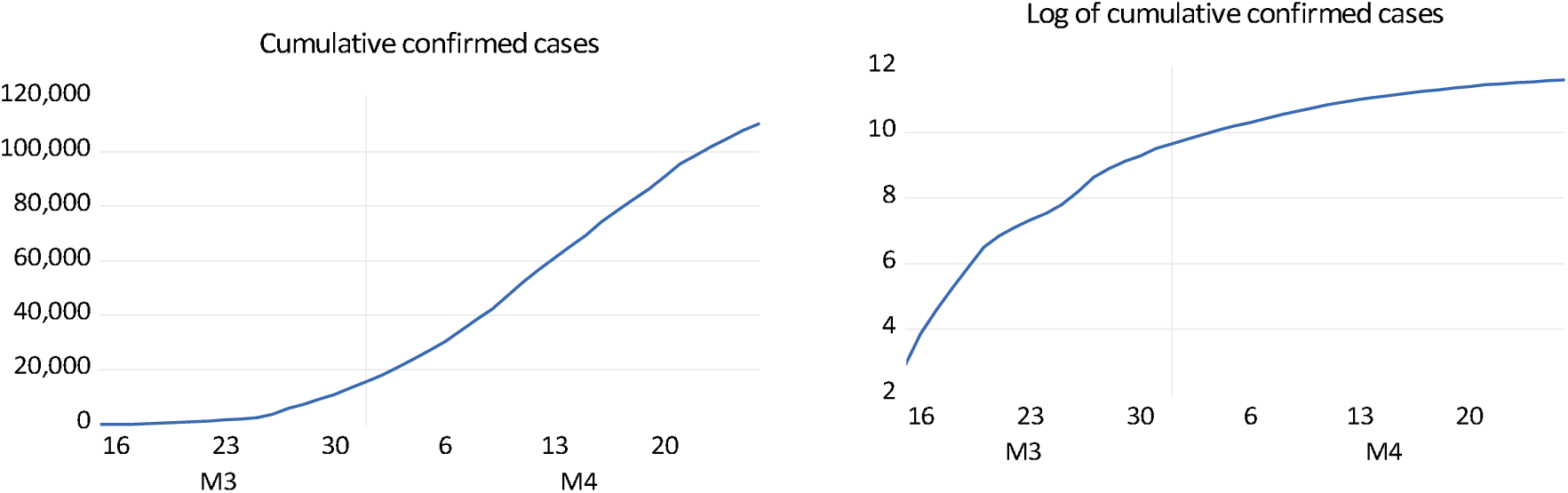
COVID-19 cumulative confirmed and log of cumulative confirmed cases The data is obtained from the Johns Hopkins University Coronavirus Resource Centre.

Officials and the general public knew that the virus was spreading around the world, they knew that they would face this problem, but they did not know when. After the first case was announced on 11 March, and with the initial rapid growth of the confirmed cases, the authorities implemented several policy measures to contain the rapid spread of Covid-19. The establishment of an independent scientific board consisting of faculty members of the medical schools and making public announcements after these committee meetings made the public follow the board’s recommendations closely. The general public started to follow the developments closely from the media and this increased public awareness of Covid-19.

Since there is a delay of approximately seven to eight days between virus infection and its reflection in statistics, the implemented policies cannot be expected to slow down the spread of Covid-19 immediately. The incubation period of the coronavirus is between 2 and 10 days, the mean estimate is 5 days (Lauer et al.; 2020; Linton et al.; 2020; WHO; 2020). Therefore, we do not expect to see the impact of the implemented policies and thus changing individual behavior on the Covid-19 confirmed cases until March 21.

When we visually examine the log confirmed cases in Figure 1, we can see that the spread of Covid-19 has slowed down from March 21onwards. Nonetheless, Covid-19 data should be analyzed by an appropriate statistical method to understand whether the decrease in the number of case’s growth rate is decreasing randomly or systematically.

## 3. Testing for intercept and trend breaks

To investigate the impact of the measures taken by the authorities, we will investigate whether there are an intercept and trend breaks in Covid-19 data.

As Hartl et al. (2020), we start with a simple log-linear trend model, but we search for unknown multiple intercept and trend breaks in the log cumulated confirmed Covid-19 cases by using breakpoint tests.

For this purpose, we specify the following simple linear trend model for log confirmed cases

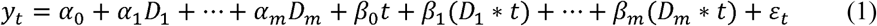

where *=:D*_1_*,…,D_m_* are dummy variables, *t* is the time trend. The residuals *ε_t_* are assumed to be normally distributed and white noise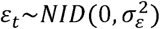.

As can be seen from Figure 1, the log confirmed cases increase quite linearly in *t* with level shifts and trend breaks, therefore, we expect specification (1) to capture the developments in the confirmed case time series well. More complex models may be required to accurately determine the drivers of log-approved cases such as weather and seasonal effects. However, these factors are currently unlikely to be correlated with a linear trend, we expect the estimates of model (1) to hardly be affected. Additionally, the time series on confirmed cases is relatively short, impeding more complex modeling of the data. We included the lagged values of the endogenous variable, but they turned out to be insignificant.

To get the growth rate of log confirmed cases, we initially estimated a model with intercept and trend by ordinary least squares, thus setting and *α*_1_ = … = *α_m_ =* 0 and*β*_1_ *=* … *= β_m_* = 0. For the simple model without intercept and trend breaks, our estimation results revealed that the intercept is 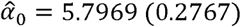 a trend coefficient is 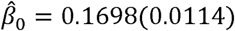, where standard errors are given in parentheses, and all parameters are significant at a 1% level. The residual standard error is estimated to be 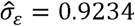. According to this result, the mean growth rate of Covid-19 confirmed cases is 17%, indicating a doubling of the confirmed cases every 4.12 days on average.

Figure 2 plots the fitted values from equation (1) without intercept and trend breaks. As can be seen from the figure, a specification without intercept and trend break initially underestimates, between 20 March and 10 April overestimates and in the last period overestimates the spread of Covid-19, which may point out level shifts and trend breaks. Following Hartl et al. (2020), we tested for a structural break in intercept and trend. However, we allowed for more than one unknown trend and intercept break. To determine the breaking point, we employed multiple unknown breakpoint tests developed by Bai and Perron (1998, 2003a, 2003b) and Yao (1988) information criterion based multiple breakpoint tests. Bai and Perron (1998, 2003a, 2003b) test is robust to serial correlation and heteroscedasticity. We adopted a general to specific methodology to determine the exact number of the break, i.e. we dropped the insignificant intercept and trend break variables from the selected model.

**Figure 2:**
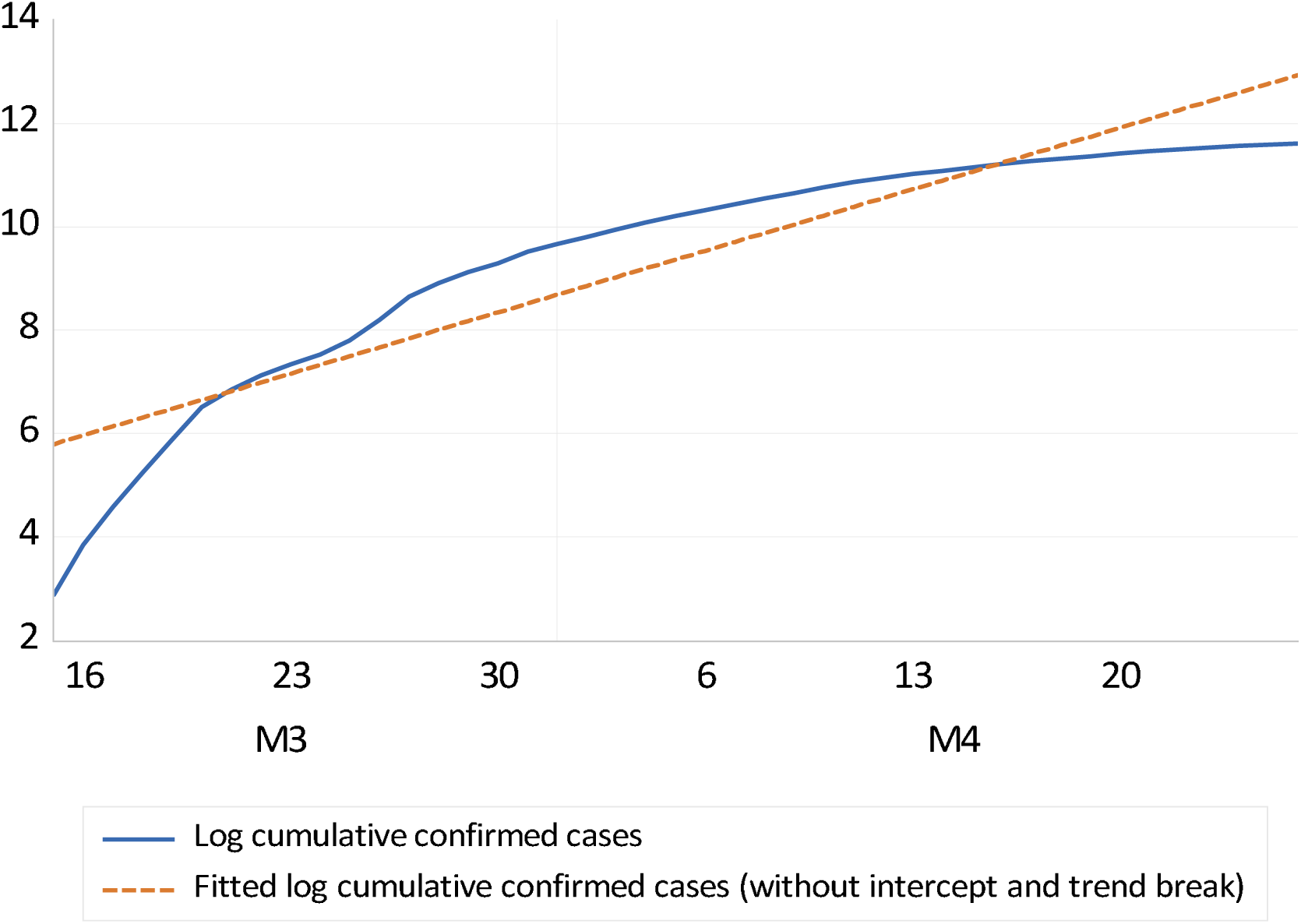
Fitted values for model (1) without intercept and trend breaks. The data is obtained from the Johns Hopkins University Coronavirus Resource Centre.

Our test results revealed that there are five potential breaks in both intercept and trend, 21 March, 27 March, 2 April, 12 April, and 20 April. The first break on 21 March reflects the initial impact of the measures, the rest of the breaks seem to reflect the growing impact of the different policy measures that are beginning to kick. We specified alternative break dates both in the intercept and trend, all other possible intercept and trend breakpoints perform substantially worse in terms of getting white noise residuals and fit.

More simply, we estimated model (1’) to determine whether measures are taken by Turkish authorities

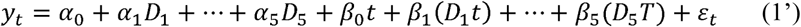

The estimation results with intercept and trend break with 2 and 5 are given in Table 1.

**Table 1:** Estimation results with 2 and 5 breaks

|  | Model 1<br>(with 2 breaks) | Model 2<br>(with 5 breaks) |
| --- | --- | --- |
| Intercept | 3.014611<br>(0.096384) | 3.052963<br>(0.038164) |
| Dum_March.21 | 2.241993<br>(0.142509) | 2.245452<br>(0.115758) |
| Dum_March.27 |  | 0.979906<br>(0.214042) |
| Dum_April.02 |  | 1.468126<br>(0.226300) |
| Dum_April.03 | 3.532695<br>(0.168484) |  |
| DUM_April.12 |  | 1.537066<br>(0.288758) |
| Dum_April.20 |  | 1.023699<br>(0.466353) |
| Trend | 0.710039<br>(0.030449) | 0.710039<br>(0.012605) |
| Trend*Dum_March.21 | -0.447516<br>(0.031773) | -0.454290<br>(0.017827) |
| Trend*Dum_March.27 |  | -0.054842<br>(0.017827) |
| Trend*Dum_April.02 |  | -0.084656<br>(0.013878) |
| Trend*Dum_April.03 | -0.191489<br>(0.010074) |  |
| Trend*Dum_April.12 |  | -0.056319<br>(0.009995) |
| Trend*Dum_April.20 |  | -0.028708<br>(0.012865) |
| Breusch-Godfrey serial correlation LM test | 29.70544$\chi^2(2)$, Prob. 0.0000 | 0.0188$\chi^2(2)$ Prob. 0.891 |
| White heteroscedasticity test | 9.444408$\chi^2(5)$, Prob. 0.0926 | 12.6288$\chi^2(9)$ Prob. 0.082 |
| Jarque-Bera test | 1.8728 Prob 0.3920 | 1.754 Prob 0.416 |
| Akaike info criterion | -1.221050 | -2.819279 |
| Schwarz criterion | -0.975302 | -2.374894 |
| Hannan-Quinn criterion | -1.130426 | -2.665878 |
| Adjusted R-square | 0.97514 | 0.98481 |
| S.E. of regression | 0.089987 | 0.052547 |

Table 1, Column 2 presents the result with two breakpoints and column 3 with five breakpoints. Information criteria indicate that the model with 5 breaks is better than the model with 2 breaks. Furthermore, the model with 2 breaks has the serial correlation in the residuals. Furthermore, Figures 3 and 4 show the log of confirmed cases and the fitted values from the model with 2 breaks and 5 breaks, respectively. It is obvious from these figures that the model with 5 breaks fits better than the model with 2 breaks.

**Figure 3:**
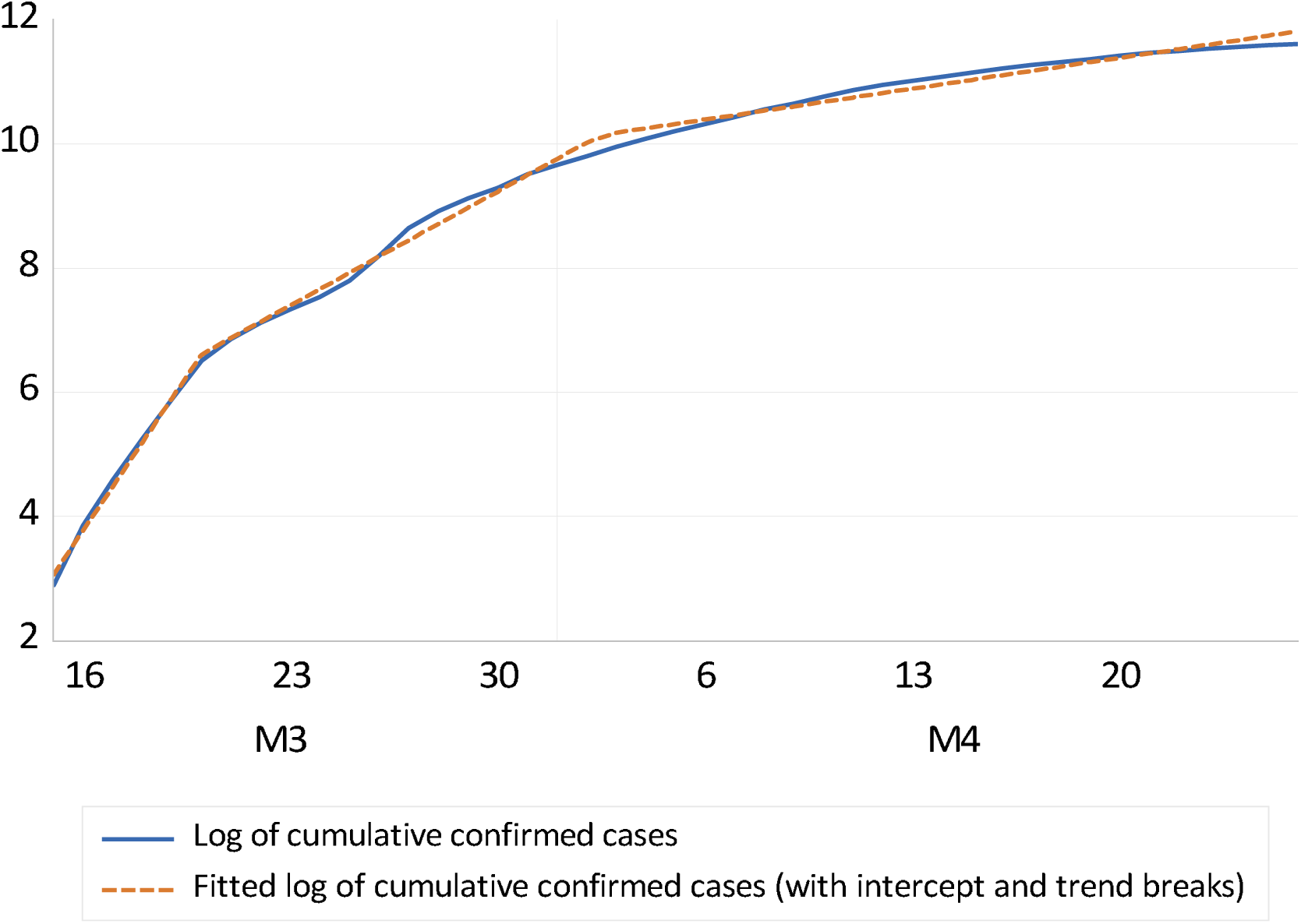
Actual and fitted values from the model (1’) with 2 breaks Source: Data obtained from the Johns Hopkins Coronavirus Resource Centre and the author’s own estimation.

**Figure 4:**
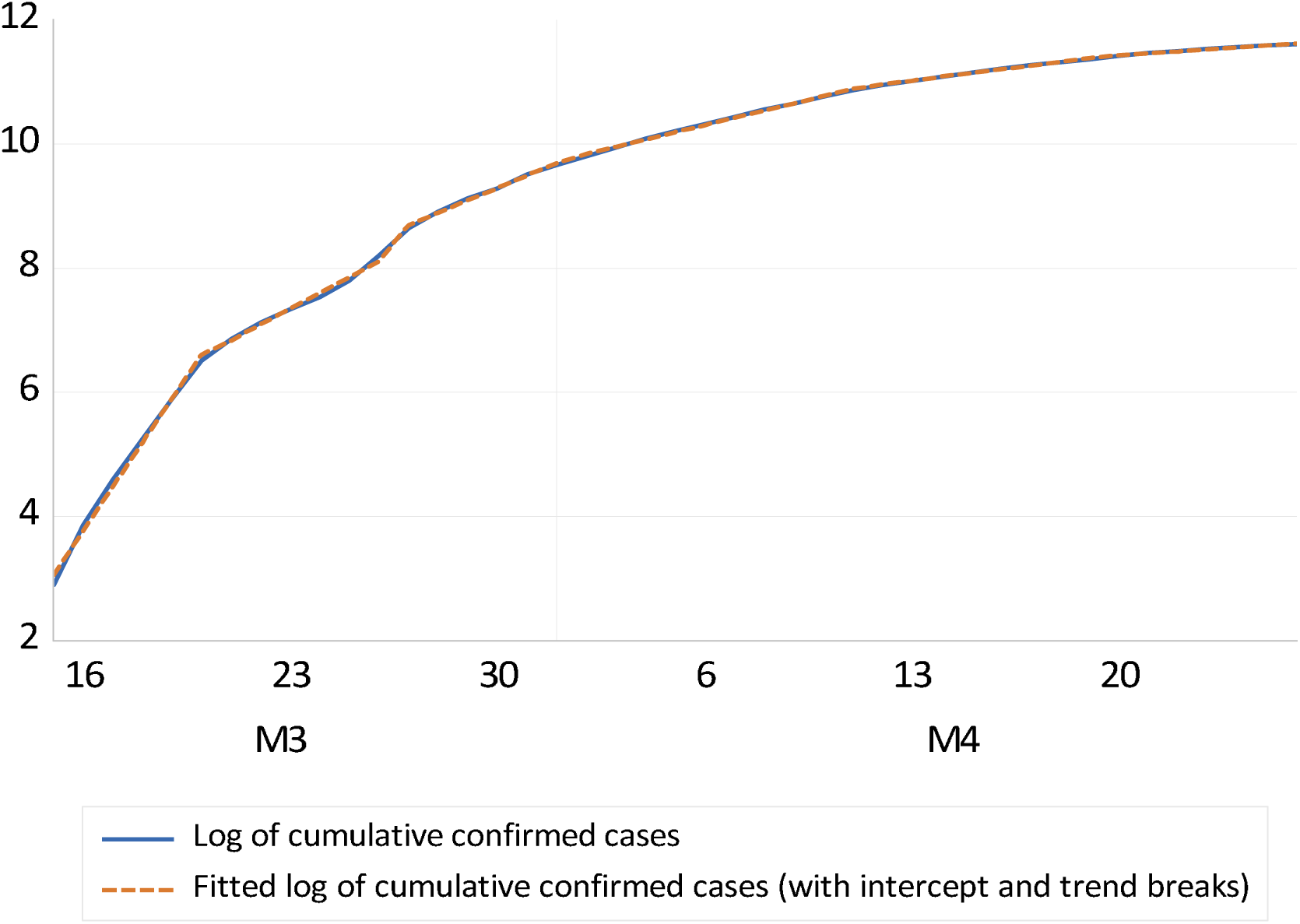
Actual and fitted values from the model (1’) with 5 breaks Source: Data obtained from the Johns Hopkins Coronavirus Resource Centre and the author’s own estimation

Therefore, we interpret the estimation results of the model with 5 breaks. All parameters are significant at a 1% level. Breusch-Godfrey serial correlation, LM test, and White heteroscedasticity test results indicate that there is no serial correlation and heteroscedasticity in the residuals, Furthermore, the Jarque-Bera test result shows that the residuals have normal distribution.

The slope estimates can be interpreted as follows. From 15 March to 20 March, the estimated daily growth rate is 71%, indicating a doubling of confirmed cases every 0.98 days. From 20 March to 27 March, the daily growth rate reduces to 25.6%, which implies a doubling of confirmed cases every 2.73 days. We can see this as an initial shocking announcement effect of the first confirmed case and governments’ a series of measures for social distancing on 12 March. From 27 March to 1 April, the daily growth reduces to 21.1%, between 1 and12 April the growth rate reduces further to 11.9%, and finally between 12 and 18 April the growth rate reduces to 6.2% which implies a doubling of confirmed cases every 11.29 days. Our results show that the growth of confirmed cases slowed considerably from 21 March onwards.

## 4. Conclusion

Our analysis confirms a pronounced slowdown in the growth of confirmed Covid-19 infections in Turkey starting from 21 March 2020. While the growth rate has slowed down significantly, the number of cases is still doubling approximately every 11 days during the last week of April. Due to substantial delays between new infections and their measurement in statistics, we could see further effects from the government measures in the near future. Corona measures need to be taken carefully and timely. During the implementation of these measures, it is necessary to avoid practices that will cause the general public to panic. If this cannot be done, social distance measures will be neglected and measures to prevent the spread of the disease will be harmed.

The spread of Covid-19 is central to the public health and the economy, therefore, we will follow the development of the confirmed growth rates closely.

## Data Availability

The data is available from Coronavirus COVID-19 global cases by the centre for systems science and engineering (https://coronavirus.jhu.edu/map.html)

## Notes

### Competing Interest Statement

The authors have declared no competing interest.

### Funding Statement

No external funding

